# Standards for Health Promoting Hospitals and Health Services: Development and tools for implementation and measurement

**DOI:** 10.1101/2024.07.22.24309820

**Authors:** Oliver Groene, Keriin Katsaros, Antonio Chiarenza, Sally Fawkes, Margareta Kristenson

**Affiliations:** University of Witten/Herdecke, Faculty of Management, Economics and Society, Germany; OptiMedis AG; Hamburg, Germany; Climate Service Center Germany (GERICS), Helmholtz-Zentrum Hereon; Department of Global Health, Institute of Public Health and Nursing Research, University of Bremen; Department of Biomedical, Metabolic and Neural Sciences, University of Modena and Reggio Emilia; La Trobe University, Department of Public Health; Department of Health, Medicine and Caring Sciences, Division of Society and Health, Linköping University

**Keywords:** health promotion, quality management, standards, self-assessment, health services, accreditation

## Abstract

The need to reorient health services towards health promotion is greater than ever. Health systems are overburdened by treating an ever-growing number of chronic patients, many of which seek care for problems that could partly be avoided or postponed through better health promotion implementation. Since its establishment, the International Network of Health Promoting Hospitals and Health Services has explicitly addressed this issue by developing specific standards on evidence-based health promotion approaches and interventions that should be implemented in health services organizations. These approaches and interventions not only address the health of patients, but also of staff and the wider community. Since the development of the standards in 2006, health systems and legitimate patient demands have evolved considerably. At the same time, topics emerged that are strongly associated with health promotion strategies, such as the climate impact of health services. An update of the 2006 standards was therefore overdue.

The purpose of this paper is, firstly, to describe the methodology used to develop and outline the 2020 Standards for Health Promoting Hospitals and Health Services. Secondly, we present a self-assessment tool, which was developed to operationalize and provide concrete measurable elements for each standard against which performance and progress towards implementation can be measured and tracked. The 2020 standards are health-oriented, continue to uphold the strategies defined in the Ottawa Charter for Health Promotion, and respond to recent international declarations and charters.

## 1. Introduction

The International Network of Health Promoting Hospitals and Health Services had its origins as a network of hospitals initiated by the European Regional Office of the World Health Organization (WHO) in the early 1990s. The network was founded on the settings approach to health promotion, as a strategy to implement the WHO’s 1986 Ottawa Charter for Health Promotion’s action area, ‘reorienting health services.’ The charter called for health care systems to contribute to the pursuit of health and argued that the “role of the health sector must move increasingly in a health promotion direction, beyond its responsibility for providing clinical and curative services “[1]. Health services need to embrace an expanded mandate which is sensitive and respects cultural needs, support the needs of individuals and communities for a healthier life, and open channels between the health sector and broader social, political, economic and physical environmental components” [1,2]. At the core of the health promoting hospitals concept was an organizational development strategy that involved reorienting governance, policy, workforce capability, structures, culture, and relationships towards the health gain of patients, staff, and population groups in communities and other settings.

To operationalize the vision and drive continuous development of health-oriented organizations, standards and complimentary self-assessment forms were formulated by WHO in 2006 [3]. The standards addressed the fundamental responsibilities of hospitals for implementing health promotion and pertained to interventions enabling health behaviors that would help prevent disease. The standards addressed leadership and managerial roles, patient assessment, information and interventions, staff health, and communication between the hospital and other care providers [3].

The 2006 standards were developed by WHO following steps proposed in ISQua’s ALPHA program, drawing on a critical appraisal of available literature and evidence, drafting, pilot testing, and implementation [4]. These original standards enabled significant international exposure and reach for the International Health Promoting Hospitals Network, as they were translated into seven languages and well-received by national health authorities, researchers, renowned scientific associations, and professional bodies.

In a next step, during the following ten year period, HPH task forces and working groups adopted this process to develop their own, domain-specific standards on equity [5], mental health [6], the environment [7], health literate organizations [8], behavioral interventions [9], patient-centered care [10], and those concerning specific population groups such as children and adolescents [11], and older adults [12]. These documents provided the basis for developing the new standards and self-assessment tool discussed in this paper.

After a decade of the 2006 standards for health promoting hospitals being applied in diverse contexts, several factors pointed towards a need to update the standards and self-assessment forms. In 2007, the hospital-focused network transitioned to become the International Health Promoting Hospitals and Health Services (HPH) Network. This change in title and scope recognized key messages from the 1978 Declaration of Alma Ata that defined the health orientation of primary health care, and called for hospitals, primary health care, and other health services to be closely linked through collaborative treatment practices, rehabilitation, health promotion, and prevention of both acute and long-term health conditions [13]. These principles were further emphasized in the 2018 Declaration of Astana launched at the WHO Global Conference on Primary Health Care and were recognized as vital to consider in an updated standards set [14].

International HPH Network members requested a revision of the standards to reflect not only the original, more comprehensive vision of health promotion in this sector, but to do so by incorporating domain-specific standards developed by International HPH Network task forces and working groups and to support and engage with several health and societal reform movements. The updated standard set was to be more concise than the sum of the existing standard sets.

At the international level, the relevance of significant developments in and affecting the health care sector needed to be accounted for. These included quality improvement, equity issues, human rights, and social movements (e.g., for women, migrants, consumers, patients, and workers), health literacy, and environmentally sustainable- “green” - health care. Strategies for empowerment were being introduced to health services through approaches such as shared decision-making and self-management support. Health care leaders were being called on to strengthen their actions and commitment towards health promotion, prevention, and societal wellbeing through global initiatives such as the Shanghai Declaration on promoting health in the 2030 Agenda for Sustainable Development [15,16]. A continuing shift in disease patterns towards non-communicable diseases was increasing the relevance of health promotion and disease prevention in countries of all levels of economic development. High-level discussions on universal health coverage (UHC) and the United Nations Sustainable Development Goals (SDGs) recognized a broader role for health care organizations to advance action to achieve societal and ecological goals [16].

A revived set of standards and self-assessment tools were developed to reflect a new, broader vision of HPH and to support hospitals and health services in mirroring the attributes reflected in the revised definition of the network:

> *Health promoting hospitals and health services (HPH) orient their governance models, structures, processes and culture to optimize health gains of patients, staff and populations served and to support sustainable societies [17]*.

The process of critically reviewing the existing standards and developing a new set commenced in 2019 with an extensive mapping analysis that identified commonalities and differences across the 2006 HPH standards and domain-specific standards. These main domains and their subdomains were identified and named the “umbrella standards” [18].

The purpose of this paper is, firstly, to describe the methodology used to develop and to outline the resulting updated standards. Secondly, the self-assessment tool is presented, which was developed to operationalize and provide concrete measurable elements for each standard against which performance and progress towards implementation can be measured and tracked.

## 2. Methods

This study was conducted in two phases. The first phase identified the 2020 Standards for Health Promoting Hospitals & Health Services and the second phase identified measurable elements for each standard, which were used to create a self-assessment tool.

### 2.1 Development of the 2020 Standards for Health Promoting Hospitals & Health Services

In early 2020, a working group led by the International HPH Secretariat was established to further refine the domains identified in the umbrella standards. A two-stage Delphi study assessed the domains using the RUMBA principles: relevant, understandable, measurable, behavioral, and actionable.

#### 2.1.1 Data Collection and Analysis

An international, multi-disciplinary expert panel comprised of International HPH Network Governance Board members, standing observers, coordinators of national and regional HPH networks, task force and working group leaders, and external subject experts (n=39) were invited via email to participate in two web-based Delphi consultations using an online survey tool. There are no ethical considerations in this study concerning data collection because no sensitive or patient data were used. All data were kept anonymous for Delphi panelists.

For both Delphi consultation rounds, the expert panel was encouraged to provide responses only to those questions which focused on topics with which they felt most comfortable. In addition to both quantitative assessments, both rounds elicited qualitative comments to help structure, align, and formulate the responses. The same set of instructions were provided to each participant.

In the first Delphi consultation round, an anonymous, quantitative survey employing a seven-point Likert scale elicited a high-level assessment of the clarity, relevance, and importance of the overarching standard dimensions and sub-standards. Mean scores were calculated for each and compiled into a tabular overview, allowing the working group to identify the most relevant and important domains. Mean scores of 5.4 and below were noted to have low clarity, relevance, and importance; mean scores of 6.5 and more were noted to have high clarity, relevance, and importance. Together with qualitative comments provided by the expert panel, the working group used these scores to condense, reorder, and regroup the standard dimensions and their components. A consolidated version of the domains and subdomains was presented for validity testing during a multi-national seminar of the International HPH Network members in the Baltic Sea region. Countries represented during the validity test included Denmark, Estonia, Finland, Germany, Norway, Poland, and Sweden. Using feedback, results were further refined based on frequency of response, and used to set up a final assessment.

In a second Delphi consultation round, quantitative ratings were made on the clarity of formulation and priority of the standard statements. Mean scores of ratings (utilizing a seven-point Likert scale) were again compiled and reviewed by the working group along with qualitative comments from the expert panel. Mean scores of 5.4 and below were noted to have low clarity, relevance, and importance; mean scores of 6.5 and more were noted to have high clarity, relevance, and importance. After identifying priority domains and subdomains and using clarity and priority scores to improve formulation, the final set of standards was produced and named the *2020 Standards for Health Promoting Hospitals and Health Services*.

### 2.2 Identification of measurable elements and development of a self-assessment tool

Based on the 2020 Standards for Health Promoting Hospitals and Health Services, a self-assessment tool was developed to operationalize the standards and to provide concrete measurable elements against which performance can be measured. Two rounds of consultation were completed to identify the measurable elements and a tool was created using feedback.

#### 2.2.1 Data Collection and Analysis

The development of the self-assessment tool began with inviting the same expert panel to propose measurable elements for each of the 86 standard statements. The expert panel was instructed to provide concise responses only to those standards which focused on topics with which they felt most comfortable. It was established that measurable elements be directly observable; able to be observed as being met or not met; based on existing documentation where possible, rather than requiring a survey; logical; widely applicable in various institutional and regional contexts; and focused on facts, documents, or other sources that help measure, observe, or prove the implementation of the standard. All participants were provided with these criteria, which were further used to assess feedback.

Responses were received and compiled in a tabular overview. A working group composed of members from the International HPH Secretariat and the German HPH Network convened to review and synthesize the responses based on frequency and adherence to the established assessment criteria. A critical review by the working group resulted in an initial list of proposed measurable elements. This was redistributed to the expert panel for their definitive evaluation of clarity and applicability of the proposed measurable element. In the last step, the internal working group incorporated feedback and produced a final list of measurable elements compiled in the *Self-Assessment Tool for implementing the 2020 Standards for Health Promoting Hospitals and Health Services*.

## 3. Results

### 3.1 The 2020 Standards for Health Promoting Hospitals and Health Services

Representatives from 14 countries participated in the Delphi panel, including the following countries/regions: Australia, Austria, Canada, Czech Republic, France, Germany, Iran, Israel, Italy, Poland, Spain, Sweden, Taiwan, and the USA. The number of responses in Delphi Round 1 was n=16, in round 2 it was n=14. Mean ratings of the clarity, relevance, and importance of the proposed standard subdimensions ranged from 5.4 (for organizational commitment to health promotion and knowledge-based and health-oriented care and service provision) to 6.6 (monitoring health needs of the population). Table 1 presents an overview of the quantitative feedback from Delphi round 1 for each of the dimensions and the ratings according to clarify, relevance and im-portance.

**Table 1.** Tabular overview of quantitative feedback from Delphi consultation round 1 using the Umbrella Standards, (mean scores, n=16).

| Dimension* | Subdimension | (1 = do not agree to 7= fully agree)<br>This dimension and its components are... |  |  |
| --- | --- | --- | --- | --- |
|  |  | ... clearly defined | ...relevant to HPH implementation | ...important for HPH implementation |
| 1: Organizational commitment | • policy and leadership<br>• measurements, including self-reported outcome, for improvement and performance | 6.2 | 5.4 | 6.6 |
| 2: Monitoring health needs for the population and patients | • the population at large<br>• service users | 6.7 | 5.9 | 6.7 |
| 3: Health of staff and the workforce | • Staff recruitment and career development<br>• Staff competencies<br>• Staff involvement<br>• Workforce health promotion and well-being | 5.6 | 5.6 | 5.6 |
| 4. Access to the service | • Entitlements/ rights to care<br>• Information to facilitate access<br>• Physical and geographical accessibility<br>• Socio-cultural acceptability | 6.4 | 5.9 | 6.5 |
| 5. Knowledge-based and health-orientated care and service provision | • Responsiveness to care needs | 6.4 | 5.4 | 6.4 |
|  | • Responsiveness to need of prevention<br>• Patient and provider communication<br>• Patient empowerment and involvement |  |  |  |
| 6. The care environment | • Respectful, trustful, and welcoming<br>• Health promoting and safe for patients<br>• Health promoting and safe for staff | 6.6 | 5.6 | 6.6 |
| 7. Participation and involvement | • Service users' engagement and impact<br>• Community engagement and impact | 6.4 | 5.9 | 6.4 |
| 8. Promoting health in the wider society | • Sharing knowledge, research, and capacity building<br>• Networking and collaboration<br>• Proactive initiatives directed to population and communities | 6.6 | 5.8 | 6.6 |

\* From the Umbrella Standards.

In addition to the quantitative feedback, respondents were asked to provide qualitative (written) feedback with suggestions for the structure and focus of the health promotion standards. Examples of this feedback are summarized in Supplementary Table 1 (a full transcript of the comments cannot be shared as it may identify some respondents).

In the second Delphi consultation round, mean ratings on the clarity of formation and priority of the proposed standard statements ranged from 4.6 for the clarity of dimension 3: Improving the health of all staff (subdimension: staff recruitment and development, umbrella standard 1) to 6.9 for the priority of dimension 3 (subdimension: staff health promotion, umbrella standard 2). Supplementary Table 2 (online only) shows each of the dimensions, subdimensions and umbrella standard statements, and their mean ratings for clarity and priority.

Qualitative comments from 14 expert panel members were likewise collected for Delphi round 2. Feedback was grouped inductively, producing the following twelve themes: positive comments, connection to HPH, health promotion, implementation, theme presentation, clarity/scope, missing topics, health workforce, patients, community, children, and language and terminology. Examples of this feedback are summarized in Supplementary Table 3 (a full transcript of the comments cannot be shared as it may identify some respondents).

Qualitative feedback from the respondents helped to reflect on the structure of the standard, formulations of specific health promotion actions, and the scope (type of organizations and cooperation partners covered). To illustrate, one response given was, “align with new HPH definition and strategy; health orientation: Implement mission of HPH not standards.” This comment was particularly useful to link the standards back to established International HPH Network steering documents and to ensure consistent terminology is used throughout the final standard set. Multiple comments highlighted the importance of “involvement of staff” in decision making processes, a vital component of a health promoting setting.

Feedback further facilitated the formulation of standards that are applicable in the diverse settings present in the International HPH Network. Given the large variability in the ratings of the health promotion dimensions and the critical feedback provided via qualitative comments, the dimensional structure of the standards, as well as their subdomains, were substantially revised. As a result, five overarching standard domains with defined objectives, 18 substandards, and 86 standard statements were determined. This represents a significant reduction in the total number of standard domains and represents widespread themes that were present across existing standards documents. Table 2 illustrates the total number of standard domains, substandards, and standard statements present prior to the Umbrella Standards and the two Delphi rounds in this study and after, resulting from the development process.

**Table 2.** Number of standards, substandards and measurable elements before and after the Delphi and feedback rounds.

| Document | Standard domain (#) | Substandards (#) | Measurable elements (#) |
| --- | --- | --- | --- |
| 2006 HPH Standards | 5 | 13 | 40 |
| Behavioral standards | 5 | 12 | 45 |
| Equity standards | 5 | 18 | 50 |
| Mental health standards | 5 | 26 | 87 |
| Environment standards | 8 | 8 | 86 |
| Patient-centered care recommendations | 3 | 3 | 24 |
| Smoke-free hospitals standards | 10 | 15 | 44 |
| Age-friendly standards | 4 | 11 | 60 |
| Standards for health literate organizations | 8 | 21 | 158 |
| Children & adolescents A | 6 | 18 | 61 |
| Children & adolescents B | 5 | 16 | 12 |
| Totals | **<u>64</u>** | **<u>161</u>** | **<u>667</u>** |
| **2020 HPH Standard** |  |  |  |
| Demonstrating organizational commitment for HPH | 1 | 3 | 13 |
| Ensuring access to the service | 1 | 3 | 11 |
| Enhancing people-centered health care and user involvement | 1 | 6 | 30 |
| Creating a healthy workplace and healthy setting | 1 | 2 | 13 |
| Promoting health in the wider society | 1 | 4 | 19 |
| Totals | **<u>5</u>** | **<u>18</u>** | **<u>86</u>** |

Domains, objectives and substandards of the 2020 Standards for Health Promoting Hospitals and Health Services are presented in Table 3. The full standards document with translations in eleven different languages are available publicly online [19].

**Table 3.** 2020 Standards, objectives, and substandards.

| 2020 Standard | Objective | Substandards |
| --- | --- | --- |
| 1: Demonstrating organizational commitment for HPH | The organization is committed to orient their governance models, policies, structures, processes, and culture to optimize health gains of patients, staff and populations served and to support sustainable societies | 1.1: Leadership<br>1.2: Policy<br>1.3: Monitoring, implementation, and evaluation |
| 2: Ensuring access to the service | The organization implements measures to ensure availability, accessibility, and acceptability of its facilities. | 2.1: Entitlement and availability<br>2.2: Information and access<br>2.3: Socio-cultural acceptability |
| 3: Enhancing people-centered health care and user involvement | The organization strives for the best possible patient-centered care and health outcomes and enables service users/communities to participate and contribute to its activities | 3.1: Responsiveness to care needs<br>3.2: Responsive care practice<br>3.3: Patient and provider communication<br>3.4: Supporting patient behavioral change and patient empowerment<br>3.5: Involving patients, families, caregivers, and the community<br>3.6: Collaborating with care providers |
| 4: Creating a healthy workplace and healthy setting | The organization develops a health promoting workplace and strives to become a health promoting setting to improve the health of all patients, relatives, staff, support workers, and volunteers. | 4.1: Staff health needs, involvement, and health promotion<br>4.2: Healthy Setting |
| 5: Promoting health in the wider society | The organization accepts responsibility to promote health in the local community and for the population served. | 5.1: Health needs of the population<br>5.2: Addressing community health<br>5.3: Environmental health<br>5.4: Sharing information, research, and capacity |

### 3.2 Identification of measurable elements and development of a self-assessment tool

The same expert panel who participated in the Delphi consultation to define the 2020 Standards proposed measurable elements for each of the 86 standard statements. The number of responses to the consultation on the measurable elements was n=13 in the first round, and n=12 in the second. Table 4 presents examples of qualitative feedback received during two consultation rounds. The rightmost column of Table 4 contains the final measurable elements, which meet the established criteria of the working group: directly observable; able to be observed as being met or not met; based on existing documentation, rather than requiring a survey; logical; widely applicable in various institutional and regional contexts; and focused on facts, documents, or other sources that help measure, observe, or prove the implementation of the standard.

**Table 4.** Example of proposed measurable elements for standard statements 1.2.1 and 1.2.3, after consultation rounds 1 and 2.

| Standard Statement | Round 1: Expert panel, proposed measurable elements | Measurable element proposed by working group | Round 2: Expert panel, final comments | Final measurable element |
| --- | --- | --- | --- | --- |
| 1.2.1 Our organization's stated aims and mission are aligned with the HPH vision. | • Submit mission and aims statements<br>• HPH principles and/or standards included in key organizational strategic documents | Our organization's mission and aims statements reflect the HPH vision. | • Presence of documented aims aligned with the HPH vision<br>• Verification of mission and vision declaration on the website | Mission and aims statements support the reorientation of hospital/health services to optimize health gains (HPH logo present, HPH website lined to homepage). |
| 1.2.3 Our organization ensures the availability of the necessary infrastructure, including resources, space, and equipment, to implement the HPH vision. | • Field observation and interviewing staff and visitors<br>• Presence of identifiable coworkers, resources, space and equipment - yes/partly/no | Through field observations and interviews we have created a list of observable elements reflecting necessary resources, space, and equipment to implement the HPH vision. | • HPH included in budget<br>• We have allocated a separate budget for the HPH project; There is a allocated budget for HPH/GNTH activities in your organisation? | A budget is designated for the implementation of HPH actions and field observations (observable elements reflecting necessary resources, staff, space, and equipment). |

A self-assessment tool was created using the final list of measurable elements proposed for each standard statement [20,21]. The tool, available as a .pdf and in Excel form, allows hospitals and health services to generate data by tracking and measuring the implementation of the standards in their organizations. The .pdf self-assessment tool allows a user to rate the implementation of each standard from a scale of 1, “not implemented,” to 10, “fully implemented.” Users can check a box for standards that are not applicable in their hospital or health service and record notes and observations after each substandard. Recorded answers can be saved so that the document can be revisited continually for reevaluation (Figure 1). Similarly, the Microsoft Excel-based tool allows users to rate the implementation of each standard utilizing an identical 10-point rating scale. With additional functionality, the tool uses the implementation ratings to generate visuals of recorded data, allowing users to clearly visualize in which areas their organization can improve and where the most progress in implementation has been achieved. In addition to generating radar charts for each substandard using the ratings of each standard statement, the tool generates bar graph overviews for each standard using the total implementation rating score of all standard statements.

**Figure 1.**
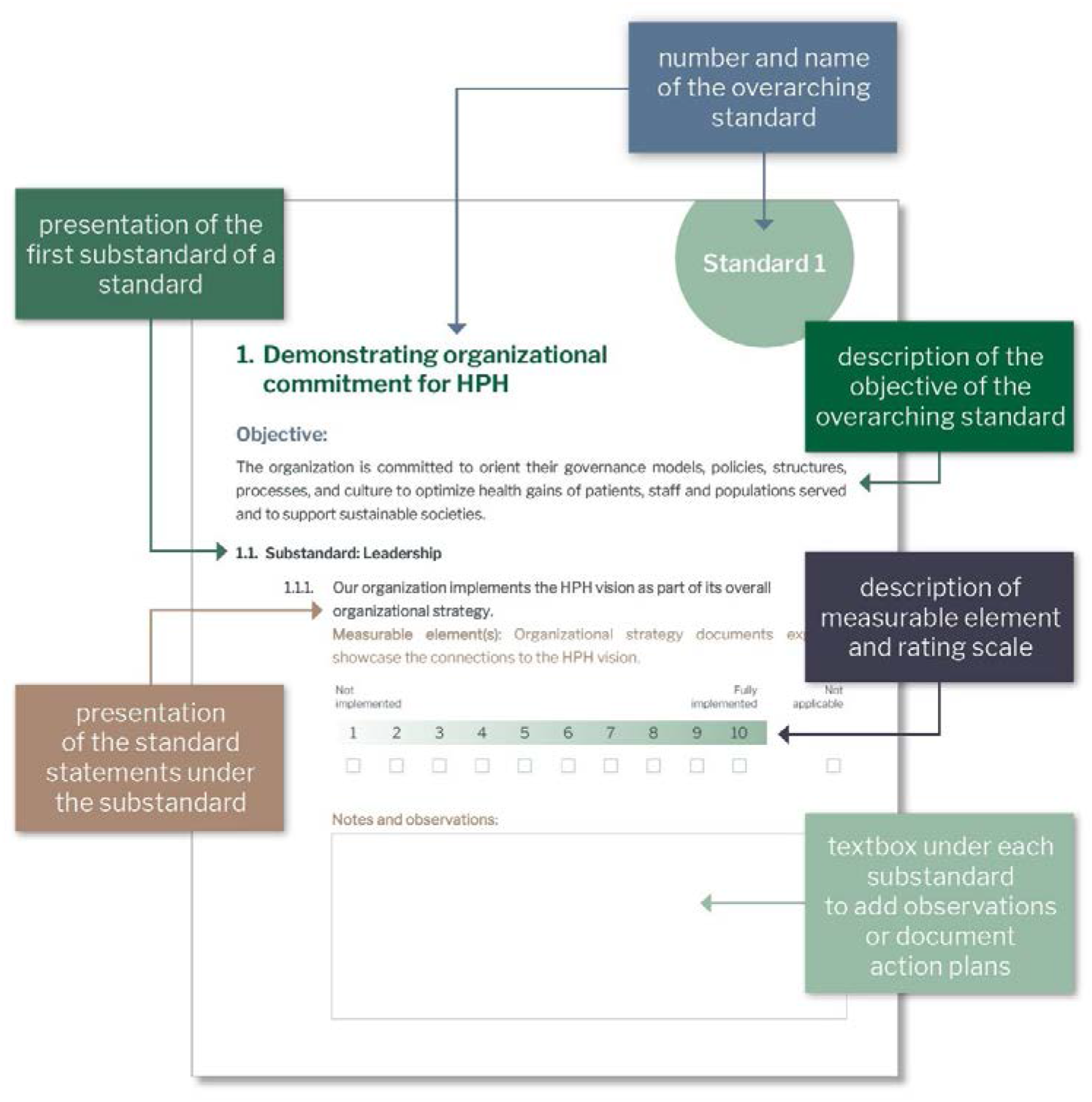
Format of the .pdf self-assessment tool.

## 4. Discussion

This paper describes the processes used to develop the 2020 Standards for Health Promoting Hospitals & Health Services and introduces the five overarching standards identified by the International HPH Network. It also describes the process used to define measurable elements for each standard which were used to develop the self-assessment tool for implementing these standards. This updating of the 2006 standards was essential for the International HPH Network. It better reflects the contemporary, comprehensive HPH vision in all facets of work, aligns with new steering documents, and better reflects the significance of health promotion within primary health care, universal health coverage, and collaborative treatment practice. Further, the updated standards address several implementation gaps that were identified with the 2006 standards.

### 4.1 Key features of the 2020 standards

Several features of the 2020 standards should be highlighted. The 2006 HPH Standards were criticized for their narrow scope, as being too focused on health behavioral interventions for patients and staff as the main strategy for improving health and having insufficient recognition of the role of environmental factors and context on health and the wider impact of health services in society [2]. To address these issues, the 2020 Standards for Health Promoting Hospitals and Health Services includes topics such as enhancing people-centered health care and service user engagement, with explicit items on empowerment strategies, creating a healthy workplace, psychosocial factors at work, and broader, more comprehensive items on the promotion of health in the wider society. Standards developed by International HPH Network task forces and working groups are included. The new standards recognize the obligations of health care organizations to society and their potential impacts on society and the health of populations. They reflect the importance of health promotion and disease prevention for tackling non-communicable diseases through primary health care strategies, universal health coverage, and collaborative treatment practices. With the inclusion of these topics, the 2020 standards have a much broader, comprehensive scope and are in harmony with the mission, values, principles, and goals brought into focus in the Global HPH Strategy 2021-2025 [22].

However, papers published on assessing the uptake of the 2006 standards referred to various implementation gaps. A study from Iran found that management and policy standards were the least implemented across the country and it was suggested that hospital and national policies should be redirected towards being more health promoting to better support the implementation of these standards [23]. Another found a lack of managerial and policy support to be a main barrier in implementing health promotion [24]. Likewise, Pelikan et. al suggest that a shift in health policy is needed to establish health promotion in the hospital and health services setting as a significant and sustainable movement [2]. A study on HPH in Taiwan credits governmental support of the HPH movement, including increased attention to organizational change, as a major reason Taiwan was able to build and maintain a large HPH network and implement numerous projects [25].

A key change made to the 2020 HPH standards offers a solution to these managerial and policy challenges. The role of leadership and governance structures was emphasized across each of the five standard domains, aligning the 2020 standards with both the Shanghai Declaration on promoting health in the 2030 Agenda for Sustainable Development and the 2030 Sustainable Development Goals. Updated standards recognize the power of those in leadership positions to influence governance and policy structures and processes in ways that would better enable commitment to implementing health promotion in their organizations.

### 4.2 Using quality management tools to facilitate implementation

The 2020 HPH Standards represent more than the value of standards in driving organizational change. In addition, assessment tools serve as a guide to operationalizing them. It has been suggested that those responsible for creating organizational support for health promotion, and implementing health promotion actions, can be motivated by tools, capacity building, and training and that insufficient leadership and a lack of evaluation procedures are barriers to implementing change [2,26]. Quality management tools, such as the 2020 HPH standards and Self-Assessment Forms, with five unique standards can be used to facilitate the implementation of health promotion in hospital and health services settings and empower relevant stakeholders to operationalize the comprehensive vision of reorienting health services set out in the Ottawa Charter for Health Promotion. Standards and tools serve as one solution to a suggested implementation gap regarding health promotion and can improve implementation and quality in hospitals and health services settings [27,28].

### 4.3 Standards and health promoting settings

The success of additional settings-based movements, such as Health Promoting Schools (HPS), can be attributed in part to the use of standards and indicators. Enforcement of the Global Standards for Health Promoting Schools, for example, has been linked to intermediate and long-term benefits for students, communities, and society [29,30]. A review of enablers and barriers of the HPS movement draws important parallels with the HPH movement. It was found, for example, that a lack of leadership and organizational support is linked to poor implementation of HPS initiatives, and that funding specified for implementing HPS is connected to development of supportive policies and uptake [31]. While these lessons underscore the value of an increased focus on management and policy in the 2020 standards, it suggests more funding needs to be dedicated to HPH by organizations, purchasing agencies and state actors.

### 4.4 Strengths and Limitations

Some limitations should be acknowledged. The expert panel selected to participate in both the development of the standards and identification of measurable elements were selected on a basis of convenience. Input from the group reflects lived experience and knowledge from only 14, mostly high income, countries. While the group included individuals from Asia, Europe, and North America, we attempted to draw out input that could be applicable across diverse national and regional contexts. Notwithstanding this, results might have been influenced by the limited number of country contexts reflected in the expert panel group. This, along with a small number of respondents (n=14) have implications for the robustness of the qualitative results in the Delphi rounds to define the standards. However, it should be noted, that in many cases individual experts consulted with teams in their organizations to provide feedback, thus the number of individuals contributing data to the Delphi rounds is much larger than our documented sample size suggests. Further adaptations of the standards and measurable elements may be needed to align the 2020 standards with quality measurement and improvement tools used in a more diverse range of contexts and to reflect other contextual factors.

As the International HPH Network grows, more information may become available from other countries, including those of varying levels of economic development. Future updates to the standards could be made to address these limitations and to reflect changes in health promotion policy and priorities.

### 4.5 HPH eCommunity

The International HPH Network has plans to launch online an “eCommunity” structured on basis of the five overarching standard domains. As of Mai 2023, the updated standards have earned acceptance by the international HPH community and have been translated from English into ten languages (Catalan, Finnish, Farsi, French, German, Hebrew, Italian, Japanese, Spanish, and Swedish) with additional translations underway. The translated documents allow for easier local implementation of the standards. The goal of the eCommunity is to provide a platform of exchange for members of HPH networks in countries/regions to link articles, tools, and videos and photos of best practice in implementing individual standards. This platform will serve as an additional capacity building tool to aid those responsible for developing favorable governance and policy conditions for health promotion in hospitals and health services and implementing health promotion action.

## 5 Conclusions

The 2020 Standards for Health Promoting Hospitals and Health Services and self-assessment tool were developed to support implementation of policy and practice, and translation of evidence, in the International HPH Network. Fourteen years of global experience and knowledge are represented in the standard sets used during the process to define this condensed set of standards. The 2020 standards are health-oriented, continue to uphold the strategies defined in the Ottawa Charter for Health Promotion, and respond to recent international declarations and charters.

This paper highlights the importance of hospitals and health services in adopting a contemporary, evidence-informed health promotion approach if they are to be credible and effective actors in improving the health of individuals and collectives represented across each of the five overarching standard domains – patients and those who support them, staff, communities, and populations. Organizations that wish to implement these standards must adopt and sustain a commitment to continuous improvement processes and proactive, innovative, and collaborative practices in their approach that are responsive to their social, environmental and health contexts.

## Supporting information

Quantitative ratings from Delphi

## Data Availability

All data produced in the present work are contained in the manuscript

## Supplementary Materials

The following supporting information can be downloaded at: www.mdpi.com/xxx/s1, Supplementary Table 1: Examples of qualitative feedback from Delphi consultation round 1 (n=16); Supplementary Table 2: Quantitative feedback from Delphi consultation round 2, (mean scores, n=14); Supplementary Table 3: Examples of qualitative feedback from Delphi consultation round 2 (n=14).

## Author Contributions

Conceptualization and methodology, O.G.; validation, O.G. and K.K.; formal analysis, O.G. and K.K.; investigation, O.G. and K.K.; data curation, O.G. and K.K.; writing—original draft preparation, O.G. and K.K.; writing—review and editing, A.C., S.F., and M.K.; supervision, O.G.; project administration, K.K.; All authors have read and agreed to the published version of the manuscript.

## Informed Consent Statement

Informed consent was obtained from all subjects involved in the study.

## Data Availability Statement

Data not containing information that could identify respondents are contained within the article or supplementary material. Data containing information that could identify respondents are not publicly available due to privacy restrictions.

---

**Supplementary Table 1:** Examples of qualitative feedback from Delphi consultation round 1 (n=16)

| Dimension and objective | Qualitative comments |
| --- | --- |
| **1: Organizational commitment**<br>The organization is committed to implement health promotion as part of their overall strategy. | The extent to which resources are made available for health promotion should be addressed, either through dedicated budgets or through [...] organizational policies.<br>You need not only the policies that leadership establishes [...] to measure performance, but identification of the structures within the organization through which the policies are operationalized. |
| **2: Monitoring health needs for the population and patients**<br>The organization collects data to identify health promotion needs and to ensure continuous improvement of health promotion efforts. | [...] I would suggest using the term healthcare systems or use subsets to populations such as community, business, healthcare system populations<br>[...] approaches may vary according to the nature of service provided and health care providers may offer multiple services. |
| **3: Health of staff and the workforce**<br>The organization ensures and improves the health of all staff, support workers and volunteers. | I would suggest using the term hospital or health system "employees." This would eliminate the perception of categories of people.<br>We suggest that this standard includes also work environment, [...] |
| **4: Access to the service**<br>The organization implements measures to ensure easy, timely and equal access to its facilities. | [...] the subdomain should be formulated in a way to favour potential contribution of the organisation in taking care of those people unable to access health promotion services because of lack of eligibility<br>This should refer to health care navigability and health literate organizations |
| **5: Knowledge-based and health-orientated care and service provision**<br>The organization strives for the best possible patient-centered care and health outcomes. | 'Patient empowerment' is a vague term with many possible interpretations and outdated. 'Patient engagement' might better express the essential aspect of a person-centered approach to care, in which the individual is actively engaged in their own care. |
| **6: The care environment**<br>The organization supports the development of a healthy, safe and respectful place for patients and staff. | An additional subdimension as "healthy and safe for the community" is suggested.<br>Would also add family/relatives, as they are also often involved in the care process and act as direct contact persons. |
| **7: Participation and involvement**<br>The organization enables service users/communities to participate and contribute to its organizational activities. | [...] engagement remains a very broad concept. If this is to make sure there is an institutional role of community and of users then it is more about looking at the mechanisms in place for such a role.<br>Involvement and participation may be interpreted very differently. Certain items needed to define the scope |
| **8: Promoting health in the wider society**<br>The organization accepts responsibility to promote health outside the health sector through cooperation and networking. | Clearer reference to environmental factors, waste and role of the hospital in the climate crisis<br>Also cooperation partners within the health sector, but in other settings may be relevant (e.g. primary care, nursing homes / long-term care, ...) |

**Supplementary Table 3:** Examples of qualitative feedback from Delphi consultation round 2 (n=14)

| Theme | Comment |
| --- | --- |
| Positive comments | The dimensions and sub-dimensions are now really clear.<br>Thanks for your hard work. It is very comprehensive. |
| Connection to HPH | They should be integrated with other HPH tools, including HPH strategy, and other HPH standards. To this end they should link assessment to improvement actions. Therefore they should be used to improve the quality of HP interventions and policies [...]. They should take into account other 'established' improvement practices in the healthcare organisations and foster their integration.<br>Align with new HPH definition and strategy; health orientation: Implement mission of HPH not standards. |
| Health promotion | They should be as 'health promoting' specific as possible.<br>Health needs, not "only" health promotion needs. |
| Implementation | They are comprehensive and relevant for HP health services. However, feasibility should be considered (they require a lot of time for the self-assessment).<br>In addition it is important to always align standards to country's rules and regulations especially when taking into consideration cultural differences. |
| Theme presentation | [...] Interesting to see, how the HLO dimensions have been merged with the HPH umbrella standards. Sometimes, different aspects are put into one standard (e.g., patients and staff) which makes it hard to tell to what extent a standard is fulfilled. Maybe better separate those in distinct items.<br>The different topics should be separated so that the standard is crystal clear what one topic the standard relates to. |
| Clarity/scope | On the level of items (standards?) the degree of detail is heterogeneous, some topics very detailed (environment), others vague.<br>Issues of equity could be more stratified – children's rights are often addressed, needs of older or vulnerable persons might need more detailed mention, perhaps also socio-cultural aspects. |
| Missing topics | No specific standard for pandemics; other crises may emerge.<br>There should be more standards on the qualities of a health literate organizations. |
| Health workforce | Staff involvement: Our organization ensures the involvement of staff in decisions impacting on the staff's working environment, as organizational and relational level as well as physical level.<br>Staff health promotion: Our organization assesses staff needs and offers health promotion concerning tobacco, alcohol, diet/nutrition, physical inactivity and stress management. |
| Patients | Supporting patient behavioural change: [...] concerning major risk factors, such as tobacco, alcohol, diet/nutrition, physical inactivity and stress management. |
| Community | Not only old volunteers but all civic society.<br>Not merely inform people but also politicians and also active collaboration with local actors. |
| Children | Subdimension "Responsive care practice" – "...management of pain and palliative care" – why limited to children; "...guidelines on high-risk screening..." – why limited to seniors? In our organization children and adolescents can ask questions freely. – why only children and adolescents? "...sharing activities in the international and/or national network..."<br>Use general standards, those suggested for children are very relevant for all patients. |
| Language & terminology | "Improvement" – reasonable if there is room for improvement – if high performance is already achieved monitoring and maintenance will be enough – so "Improvement/ Maintenance [...]"<br>I propose some words (Stress management, Self-Care Skills, Lifestyle Medicine, Organizational and relational level, wellbeing recovery [...])! |

